# Gut microbiome and lichen sclerosus: a two-sample bi-directional Mendelian randomization study

**DOI:** 10.1101/2024.07.23.24310907

**Authors:** Jiayan Chen, Peiyan Wang, Changji Xiao, Kalibinuer Kelaimu, Xianshu Gao, Xiaomei Li, Jun Hu

**Affiliations:** Department of Radiation Oncology, Peking University First Hospital, Beijing, China; School of Information, University of Michigan, Ann Arbor, MI, United States; Department of Obstetrics and Gynecology, Peking University First Hospital, Beijing, China

**Keywords:** gut microbiome, lichen sclerosus, causal relationship, Mendelian randomization, genome-wide association study

## Abstract

**Background:** Recent studies suggest a potential link between gut microbiomes (GMs) and inflammatory diseases, but the role of GMs in lichen sclerosus (LS) remains unclear. This study aims to investigate the causal relationship between GMs and LS, focusing on key GM taxa.

**Methods:** We utilized GWAS summary statistics for 211 GM taxa and their association with 2,445 LS patients and 353,088 healthy controls, employing Mendelian randomization (MR). GWAS data for GM taxa came from the MiBioGen consortium, and for LS from the FinnGen consortium. The primary analytical tools included the inverse-variance weighted (IVW) method, weighted MR, simple mode, weighted median, and MR-Egger methods. Sensitivity analyses included leave-one-out analysis, MR-Egger intercept test, MR-PRESSO global test, and Cochrane’s Q-test. A reverse MR analysis was conducted on bacteria identified in the forward MR study.

**Results:** We identified one strong causal relationship: order *Burkholderiales* [odds ratio (OR) = 0.420, 95% confidence interval (CI): 0.230 - 0.765, p = 0.005], and three nominally significant relationships: phylum *Cyanobacteria* (OR = 0.585, 95% CI: 0.373 - 0.919, p = 0.020), class *Betaproteobacteria* (OR = 0.403, 95% CI: 0.189 - 0.857, p = 0.018), and genus *Butyrivibrio* (OR = 0.678, 95% CI: 0.507 - 0.907, p = 0.009). Moreover, this MR analysis was not impacted by horizontal pleiotropy, according to the MR-Egger intercept test and MR-PRESSO global test (p > 0.05). Remarkably, the reliability of our results was confirmed by leave-one-out analysis. Reverse MR analysis showed no significant causal relationship between LS and GM.

**Conclusions:** This MR study identifies specific gut flora linked to a lower risk of LS, offering new insights for disease treatment and prevention. Future research should incorporate metagenomics sequencing of extensive microbiome GWAS datasets.

## 1. Introduction

Lichen sclerosus (LS) is a chronic mucocutaneous immune-mediated disease that is characterized by skin atrophy and hypopigmentation [1]. A majority of the LS patients, exceeding 80%, present with anogenital lesions. Common Typical extragenital sites involve neck, shoulders, upper trunk, and thighs [2]. The typical clinical presentation consists of persistent white atrophic patches in the external genitalia, often accompanied by discomfort and itching. Potentially, the lesions could worsen and cause the vaginal introitus to constrict and scar, which would impair urogenital and sexual function. Furthermore, women with vulvar LS have a high risk of squamous cell carcinoma with a lifetime incidence rate ranging from 0.4% to 6% [3-4]. The prevalence of genital LS was reported to be 1.7% in gynecological practice [5]. However, the precise incidence of LS remains understated. LS can manifest at any age and has an impact on both sexes, with a prevalence ratio of females to males ranging from 3:1 to 10:1 [6]. Postmenopausal women are most commonly affected, followed by men, prepubertal children, and adolescents [7].

It is still difficult to fully comprehend the pathophysiology of LS. LS is characterized by a hereditary and familial propensity; however, chronic irritation, endocrine status, and autoimmune diseases may also contribute to the pathogenesis [8-11]. The microbiota is a key immune system modulator that contributes significantly to homeostasis maintenance. Over the past 20 years, numerous investigations have been carried out to examine the constitution and the inflammatory function of intestinal bacterial flora in preclinical and clinical studies focusing on dermatologic conditions, such as alopecia areata, rosacea, atopic dermatitis, and psoriasis [12-14]. Certain notable associations between microbiome signatures have been discovered. The relationship between LS and gut microbiota, however, is poorly understood. Nevertheless, further research is required to investigate the specific role of different GM taxa in the genesis of LS.

In epidemiological research, the Mendelian randomization (MR) method, which employs genetic variants as an instrumental variable, is widely used to investigate, the potential causal effect of exposure to specific diseases. In the MR approach, the genetic alleles could be randomly distributed like a randomized controlled trial, which is less likely to be impacted by confounding or reverse causation. Hence, we conducted a bidirectional large-scale MR analysis to identify potentially influential GM taxa using summary data from GWAS in order to shed light on the prevention and treatment of LS.

## 2. Materials and Methods

### 2.1. Study design

Figure 1 shows the design of the two-sample MR analysis. We followed three important MR assumptions to determine the possible causal association between GM and LS risk: IVs must be correlated with GM, they must be independent of confounding variables, and IVs must only affect LS risk through GM [15].

**Figure 1.**
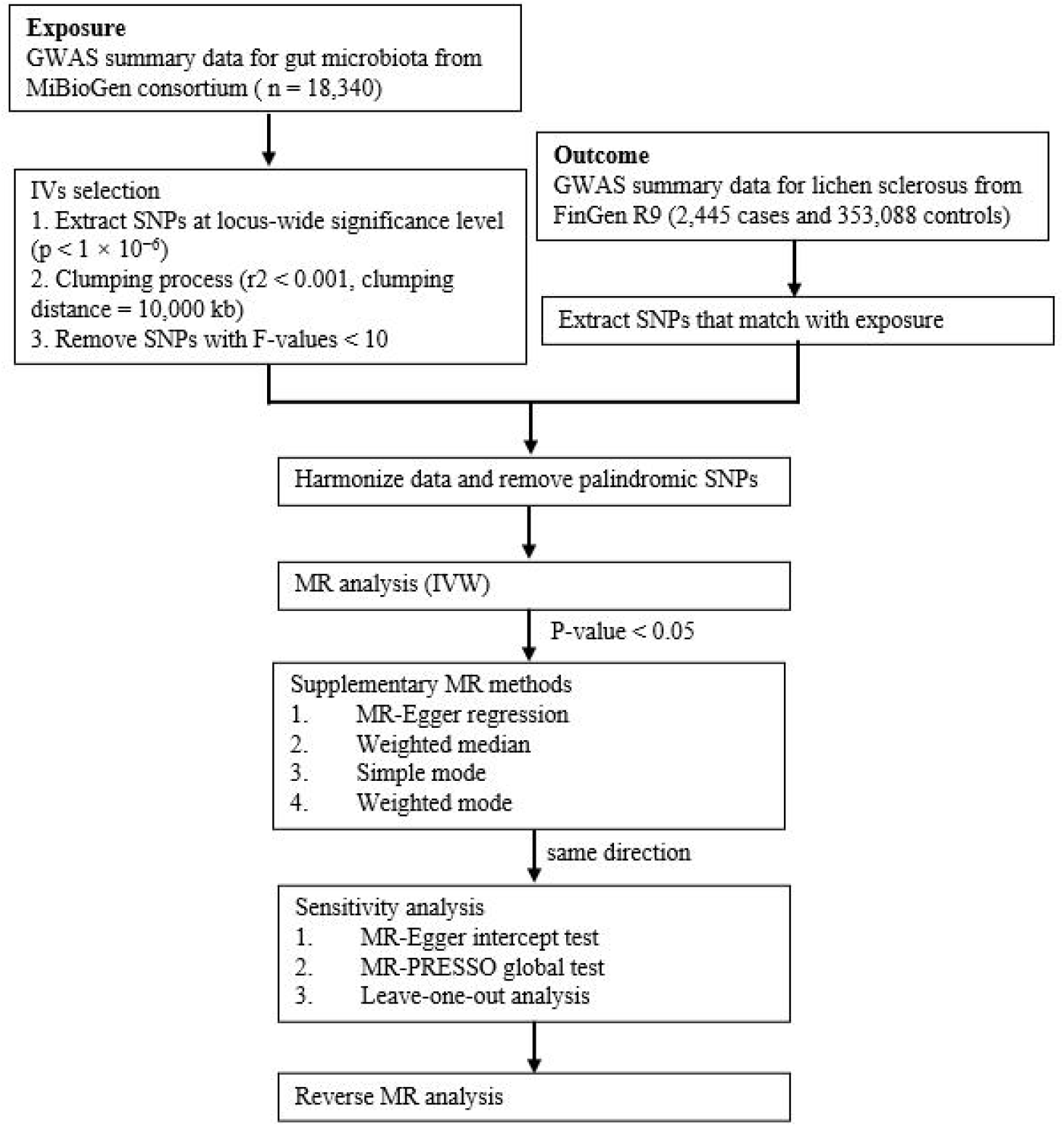
Study design and workflow.

### 2.2. Exposure data

16S fecal microbiome rRNA gene sequencing profiles and host genotypes of 18,340 participants were examined in-depth by the MiBioGen consortium [16]. A comprehensive GWAS encompassing 211 GM taxa from genus to phylum revealed genetic variations linked to 9 phyla, 16 classes, 20 orders, 35 families, and 131 genera. The GM GWAS summary statistics can be accessed by downloading them from https://mibiogen.gcc.rug.nl/ [17–19].

### 2.3. Outcome data

We acquired LS-related GWAS summary statistics from the FinnGen consortium R9 release, encompassing 2,445 cases and 353,088 controls, available at https://r9.finngen.fi/. It is noteworthy that our study, relying on publicly available databases, did not require additional ethics approval or informed consent.

### 2.4. Instrumental variable (IV) selection

In this MR study, single-nucleotide polymorphism (SNP) that exhibited strong association with each GM taxon were utilized as IVs. For a more comprehensive analysis, IVs achieving locus-wide significance (p < 1 × 10 ^−6^) were selectively employed. Concurrently, the PLINK clumping method was applied to exclude SNPs in linkage disequilibrium (r^2^ < 0.001, kb = 10,000). The degree of weak instrumental bias was then estimated using the F-statistic of IVs, where a value of >10 indicated the lack of bias resulting from weak IVs [20]. Palindromic SNPs and those not found in the outcome were finally eliminated from the instrumental variable group.

### 2.5. Statistical analysis

The IVW approach, an extension of the Wald ratio estimator based on meta-analysis principles, was the main technique used in this MR investigation to infer causality [21]. Four more MR methods—MR-Egger, weighted median, weighted mode, and simple mode—were then utilized to supplement the IVW results if the IVW technique showed a causal association (p < 0.05) for each GM taxon [22–23]. At the p-value of less than 0.05, causal relationships were shown as OR with 95% CI. False discovery rate (FDR) correction was used with a threshold of q < 0.05 to address multiple tests. Exposure-outcome couples exhibiting a consistent direction were required to demonstrate consistency across all MR techniques, demonstrating a causal link. Many sensitivity studies were carried out in order to evaluate the stability of causal associations. The horizontal pleiotropy was determined using the MR-Egger intercept test and the MR-PRESSO global test [24–25]. Moreover, result robustness was evaluated using a leave-one-out analysis. Reverse MR studies were conducted to examine if identified major bacterial genera were causally impacted by LS, employing SNPs linked with LS as IVs. The R software’s “TwoSampleMR” and “MR-PRESSO” packages (version 4.2.2) served as the foundation for all studies.

## 3. Results

### 3.1. IVs Overview

At the phylum, class, order, family, and genus levels, we found a total of 16, 26, 26, 54, and 149 SNPs linked to GM, with a significance level of p < 1 × 10 ^−6^. Notably, every IV showed stronger correlations between exposure and result (P_exposure_ < P_outcome_), and every F-value was greater than 10. Supplementary Table S1 has comprehensive information about the IVs.

### 3.2. MR Analysis

In our implementation of the Inverse Variance Weighted Fixed Effects (IVW-FE) technique to assess the causal relationship between LS and 211 GM taxa in five hierarchical levels, several taxa were identified as potentially influencing LS risk. Specifically, phylum *Cyanobacteria* (id: 1500), class *Betaproteobacteria* (id: 2867), order *Burkholderiales* (id: 2874), and genus *Butyrivibrio* (id: 1993) exhibited a decreased risk for LS (Figure 2). Following FDR correction, the protective effect of order *Burkholderiales* (id: 2874) [OR= 0.420 (0.230 - 0.765), p = 0.005, q = 0.036] against LS remained significant. Importantly, Cochran’s Q test results indicated an absence of heterogeneity.

**Figure 2.**
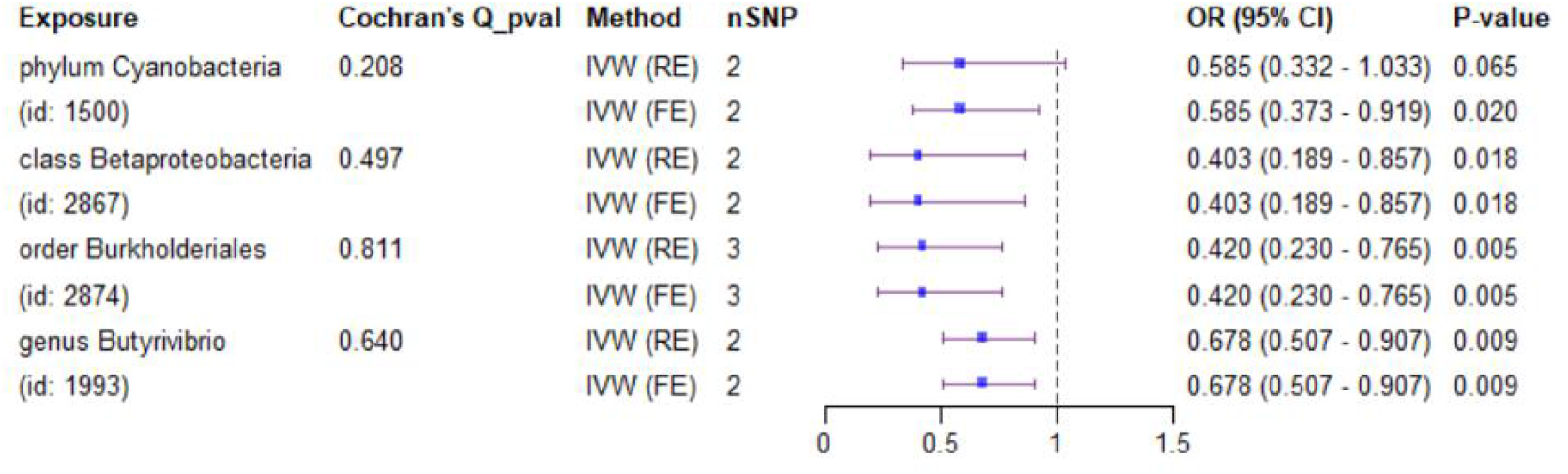
MR results of causal effects between GM and LS risk.

To further support the causal relationship between these GM taxa and LS, we utilized four more MR techniques: MR-Egger, weighted median, weighted mode, and simple mode (Figure 3). The robustness of our observed causal relationships was reinforced by the encouraging consistency of the results obtained from these approaches with the IVW findings.

**Figure 3.**
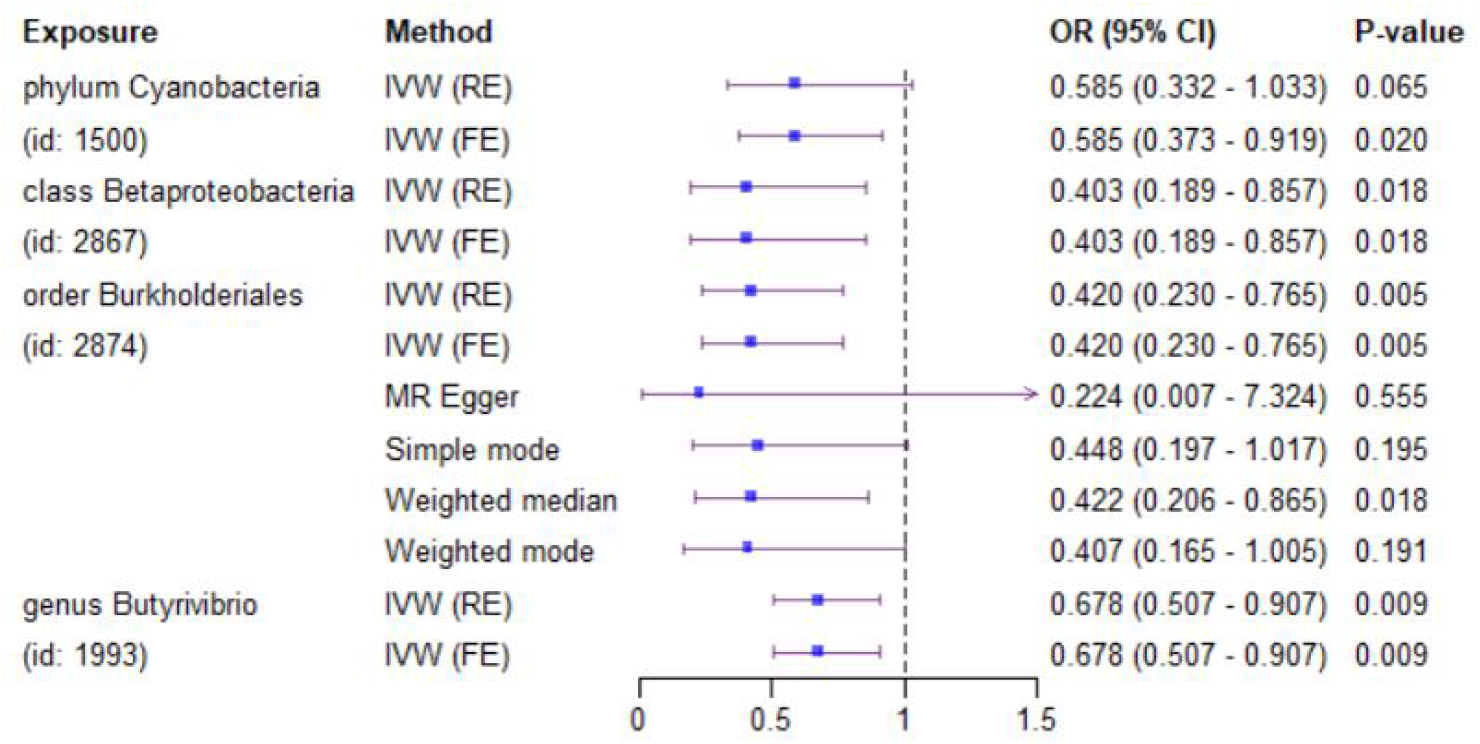
Various MR outcomes for four GM taxa with identified causal associations to LS.

### 3.3. Sensitivity analysis

The MR-Egger intercept test results revealed no horizontal pleiotropy (p > 0.05) within the IVs associated with order *Burkholderiales* (id: 2874) and LS (Supplementary Table S2). Furthermore, the leave-one-out analysis demonstrated the robustness of the MR results, as the removal of any particular IV did not change the overall findings for order *Burkholderiales* (id: 2874) (Supplementary Figure S1). It is noteworthy, however, that due to an insufficient number of SNPs, MR-Egger intercept tests and leave-one-out analyses could not be conducted for the remaining three gut microbiota taxa. Similarly, the MR-PRESSO global test was precluded for the same reason.

### 3.4. Reverse MR

Nine SNPs linked to LS were found to be eligible IVs after a thorough series of IV screening procedures (Supplementary Table S3). The findings of the reverse MR analysis, displayed in Supplementary Table S4, yielded no proof of a causal effect from LS to the determined bacterial taxa. The results of the MR-Egger intercept test and the MR-PRESSO global test both indicated that there was no horizontal pleiotropy among the remaining IVs. A comprehensive presentation of the sensitivity tests can be found in Supplementary Table S5, demonstrating the robustness of our findings in the face of various analytical challenges.

## 4. Discussion

The gut microbiota is one of the potential targets for regulating the host’s immune responses. The correlation between the composition of the gut microbiome and the immunological system regulation has provided a novel perspective on the pathogenesis of numerous chronic multifactorial diseases. According to multiple studies that link gastrointestinal homeostasis to skin disorders, both the component and function of the GM are altered in patients with autoimmune and inflammatory skin diseases. Atopic dermatitis (AD) development and progression may be influenced by alterations in the microbiota diversity. For instance, decreased variation of the GM is associated with more server disease. Recent evidence suggests that microbiota could be involved in modifying itchiness in atopic dermatitis through the interactions between the gut, skin and brain. Damage to the epidermal barrier, brain sensitization of pruritus-generating mechanisms, and modulation of histamine-independent itch are caused by the presence of proinflammatory cytokines, microbial metabolites, and a compromised immune response [26]. Microbiota dysbiosis was found to induce an irregular immune response in individuals with psoriasis, and the alterations of microbiota composition were linked to the level of inflammation-related biomarkers that were abnormal in the patients with psoriasis. For example, the relative abundances of *Phascolarctobacterium* and *Dialister* may serve as possible indicators of the psoriasis progression, as the IL-2 receptor showed a positive correlation with the former and a negative correlation with the latter [27].

However, only a few studies have shown a link between changes in GM and LS disease. Chattopadhyay et al. conducted a pilot case-control study with prepubescent females, which suggested that the females with vulvar LS exhibited an increased relative abundance of Escherichia coli, Bifidobacterium adolescentis, Akkermansia muciniphila, Clostridiales spp., Paraprevotella spp., and Dialister spp in comparison to healthy controls, while the abundance of Roseburia faecis and Ruminococcus bromii was noticeably decreased. These findings showed a potential correlation between intestinal dysbiosis and juvenile vulvar LS [28]. Nygaard et al.’s study revealed that women with LS had a higher relative abundance of phylum Euryarchaeota in their GM than controls [29]. However, the inconsistent findings and small sample of these studies pose challenges to reaching generalizable conclusions. Additionally, due to variations in gender, race, age, and body region, the composition of the GM may also differ among studies. The presence of these unclear factors impedes the ability to establish a causal relationship between GM and the risk of LS disease.

Our work thoroughly evaluated the causative influence of 211 GM taxa (from phylum to genus level) on LS, emphasizing the importance of GMs in LS. Four causal links in all were found, three of which were nominal and one of which was strong. Our MR analysis has provided the first confirmation that class *Betaproteobacteria* (id.2867), genus *Butyrivibrio* (id.1993), order *Burkholderiales* (id.2874), phylum *Cyanobacteria* (id.1500) have a protective effect LS. Most of these bacteria belong to Phylum Firmicutes and Proteobacteria, which was consistent with our previous research with vulvar skin microbiota [30]. It indicates an interesting linkage between gut and genital skin microbiota. As far as we know, this is the first MR investigation to look into the possibility of a causal relationship between gut microbiota and LS. In accordance with researches in other inflammatory diseases, our results suggest that gut dysbiosis may play a role in the development of LS.

It was believed that the metabolic activity and immunological response of gut microbiome may affect skin conditions [31]. The gut barrier integrity is primarily maintained by the microbial communities, through the conversion of polysaccharides and vitamins, as well as short-chain fatty acids. For example, butyrate reduces interstitial barrier permeability and improves the integrity of the epithelial barrier. Genus *Butyrivibrio*, class *Betaproteobacteria*, and order *Burkholderiales* are commensal bacteria that are well recognized for their capacity to generate short-chain fatty acids, particularly butyrate, which could help maintain systemic immunity balance [32]. Butyrate serves as the main source of energy for the colonic epithelium and exerts anti-inflammatory effects in the colonic mucosa by preventing the activation of NF-κB [33]. Several studies have shown that patients with Crohn’s disease and ulcerative colitis, respectively, have a decreased enrichment of *Butyrivibrio* in colon and saliva samples than healthy controls, respectively [34-35]. Additionally, lower concentrations of stool butyrate have been observed in those suffering from inflammatory bowel disease (IBD), providing more evidence to support the possibility that gut dysbiosis plays a causal role in inflammatory conditions like IBD and potentially LS. Nevertheless, the relationship between skin disease and immunological activity triggered by the gut microbiome remains obscure, and further investigation is needed.

Our work contributed substantial evidence to the current literature concerning the causal relationship between gut microbiota and LS. The primary benefits of our research are: (1) We analyzed genetic data from a large sample group, which increases the reliability of the results in comparison to smaller observational research. (2) Through MR analysis, confounding variables are eliminated from causal association, and the causal association identified in our study may serve as potential candidates for future functional investigations. This study has some limitations that should be mentioned. The primary limitation of our work is that the most of participants in our research were limited to people who had European origins. Although reducing bias caused by population heterogeneity would be achieved, further research is necessary to explore whether the MR results can be general in other populations. Second, in order to obtain a greater number of SNPs, we relaxed the p threshold, which may heighten the likelihood of contravening the basic MR design premise. However, all SNPs have F statistic greater than 10, which indicates that all weak SNPs were excluded from the MR estimation. And rigorous FDR correction was applied to identify the significant results to reduce the likelihood of false-positive findings. Third, we were unable to fully reduce pleiotropy due to the fact that the specific biological functions of the utilized SNPs are still unknown. However, it is encouraging to note that distinct MR models generated consistent estimates, and the analyses of sensitivity analyses under various assumptions were unable to identify any horizontal pleiotropy. Studies have indicated that LS patients frequently exhibit several comorbidities, such as depression and behavioral disorders. The prevalence rates of these comorbidities may reach 37% and 63%, respectively [36-37]. Many of these comorbidities have been demonstrated to have an association with the GM [38]. Therefore, it is exceedingly probable that variations in GM exist between the patients with LS alone and those with LS accompanied by other medical conditions. However, the original GWAS data failed to make the distinction, thus additional research is required to solve this limitation.

## 5. Conclusions

Our study serves as a foundation for recognizing the causal association between the gut microbiota and LS. In addition, it was found that a number of gut flora may lower the incidence of LS. These findings hold promise for their prospective use in the prevention and management of LS. This would have an implication to clinicians that microbiome-targeted therapies might be promising preventive and therapeutic tools for LS. Future investigation is required to unravel the fundamental mechanism. Additional observational studies or lab-based research is warranted to substantiate these results.

## Supporting information

Supplemental Tablels

## Data Availability

All data produced in the present study are available upon reasonable request to the authors.

https://mibiogen.gcc.rug.nl/

https://r9.finngen.fi/

## Supplementary Materials

The following supporting information can be downloaded at: www.mdpi.com/xxx/s1, Figure S1: title; Table S1: title; Video S1: title.

## Author Contributions

Jiayan Chen, Peiyan Wang, and Jun Hu designed the research. Peiyan Wang collected and analyzed the data. Xianshu Gao and Xiaomei Li offered technical advice. Jiayan Chen, Peiyan Wang, and Jun Hu drafted the manuscript. Jun Hu supervised the study. Changji Xiao and Kalibinuer Kelaimu revised the manuscript. All authors approved the submitted version of the article and participated in its writing.

## Data Availability Statement

The summary data of MiBioGen can be downloaded from the website https://mibiogen.gcc.rug.nl/. The summary data of FINNGEN can be downloaded from the website https://r9.finngen.fi/. This published work and its supplementary information files contain other datasets that were generated and/or analyzed during the current study, which are accessible to the public.

## Acknowledgments

The researchers of MiBioGen and FinnGen, as well as the members of all GWAS cohorts that were used in this study, are appreciated by the authors for providing the GWAS summary statistics.

## Disclaimer/Publisher’s Note

The statements, opinions and data contained in all publications are solely those of the individual author(s) and contributor(s) and not of MDPI and/or the editor(s). MDPI and/or the editor(s) disclaim responsibility for any injury to people or property resulting from any ideas, methods, instructions or products referred to in the content.

