## Supplemental Figures for "Gut microbiome and lichen sclerosus: a two-sample bi-directional Mendelian randomization study"

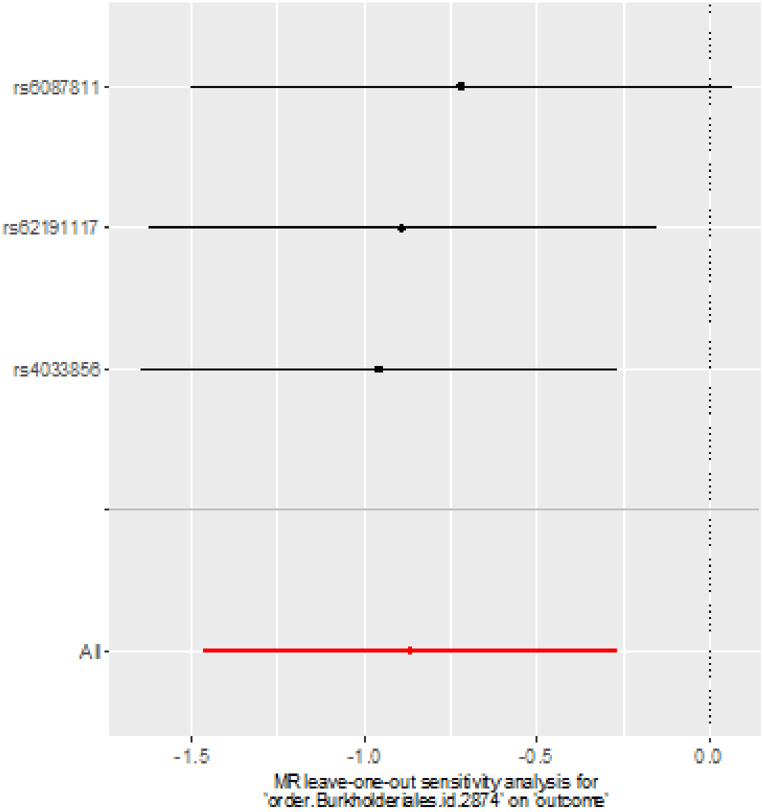


Supplementary Figure S1. Leave-one-out analysis of the causal effect of LS on order *Burkholderiales*. Red lines represent estimations from the IVW test. The number of SNPs of phylum *Cyanobacteria*, class *Betaproteobacteria*, and genus *Butyrivibrio* associated with LS is insufficient for leave-one-out analysis.
